# Vaccine hesitancy and anti-vaccination attitudes during the start of COVID-19 vaccination program: A content analysis on Twitter data

**DOI:** 10.1101/2021.05.28.21257774

**Authors:** Hüseyin Küçükali, Ömer Ataç, Ayşe Seval Palteki, Ayşe Zülal Tokaç, Osman Erol Hayran

**Author notes:** Corresponding author (HK).

## Abstract

**Background:** Vaccine hesitation, which is defined as one of the most important global health threats by World Health Organization, maintains its universal importance during the COVID-19 period. Due to the increasing appearance of anti-vaccine arguments on social media, Twitter is a useful resource in detecting these contents. In this study, we aimed to identify the prominent themes about vaccine hesitancy and refusal on social media during the COVID-19 pandemic.

**Methods:** In this qualitative study we collected Twitter contents which contain a vaccine-related keywords and published publicly between 9/12/2020 and 8/1/2021 (n=551,245). A stratified random sample (n=1041) is selected and analyzed by four researchers with content analysis method.

**Results:** All tweets included in the study were shared from 1,000 unique accounts of which 2.7% were verified and 11.3% organizational users. 90.5% of the tweets were about vaccines, 22.6% (n=213) of the tweets mentioned at least one COVID-19 vaccine name and the most frequently mentioned COVID-19 vaccine was CorronaVac (51.2%). Yet, it was mostly as “Chinese vaccine” (42.3%). 22.0% (n=207) of the tweets included at least one anti-vaccination theme. Among tweets that included an anti-vaccination theme; poor scientific processes (21.7%), conspiracy theories (16.4%), and suspicions towards manufacturers (15.5%) were the most frequently mentioned themes. The most co-occurred themes were “Poor scientific process” theme come along with “suspicion towards manufacturers” (n=9) and “suspicion towards health authorities” (n=5).

**Conclusions:** This study may be helpful for health managers to identify the major concerns of the population and organize the preventive measures, through the significant role of social media on early information about vaccine hesitancy and anti-vaccination attitudes.

## Introduction

Vaccine hesitancy has been defined by World Health Organization’s (WHO) advisory group Strategic Advisory Group of Experts on Immunization as “delay in acceptance or refusal of vaccines despite availability of vaccine services”. (1) Although vaccination is the key prevention strategy against communicable diseases there is an increasing hesitancy that is identified to be among ten threats to global health by WHO. (2) Vaccine hesitant individuals who are not homogeneous and influenced by many factors, have different attitudes ranging from some acceptance to complete refusal for different vaccines. Determinants of vaccine hesitancy have been described under three domains, contextual influences (socio-economic, cultural, political, health system factors etc.), individual and group influences (knowledge, beliefs, attitudes, experiences etc.), and vaccine/vaccination-specific issues (costs etc.) and each determinant can be a barrier or promoter factor. (1,3)

Studies related to acceptance and refusal about the COVID-19 vaccine show that although attitudes vary from population to population, hesitancy is a universal problem. Factors affecting acceptance or hesitancy have been stated as fear of side effects, safety, effectiveness, beliefs about unnecessariness, lack of information, short duration of immunity and general vaccine refusal. Another important conclusion is that the acceptance of COVID-19 vaccine rate has declined over time. (4)

In recent decades publicly posted materials on social media become valuable sources for identifying people’s beliefs and attitudes about important health issues and understanding socio-cultural context. (5) Twitter comes forward among other social media sites, as it hosts real-time sharing of emotions and contents, and provides easier access to broad populations. Data from these sources are used for content analysis, network analysis, surveillance and even interventions in the public health field. (6)

Due to its importance on public opinion, attitudes and information seeking behaviors about vaccination are also searched via social media in different ways. A study which analyzed the contents of the most frequently visited vaccine-focused blogs and forums shows that the information was more controversial and the attitude toward vaccination was more negative on non-expert moderated sites. (7)

When the sentiments of vaccine-related tweets between 2011 and 2019 are analyzed, it is seen that both positive and negative sentiments in the posts have increased in number and proportion over time compared to neutral sentiment. This increase is especially apparent during traditional media discussions about vaccines. (8) Although the number of users who produce anti-vaccine content on social media is fewer, it is also seen that they have more presence and interactions. (9) The content of anti-vaccine posts often include personal stories, negative health impacts attributed to vaccination, discussions about vaccine components, distrust of the pharmaceutical industry, criticism of vaccine research, political debates and conspiracy theories. (9, 10)

Increasing apparency and popularity of vaccine hesitant contents on social media is also a growing public health concern during the pandemic period and threatens public acceptance of newly developed vaccines against the COVID-19. (11)

In this study, we aimed to identify the prominent themes about vaccine hesitancy and refusal on social media during the COVID-19 pandemic.

## Material and methods

This is a qualitative study with content analysis design. Turkey imported Sinovac’s vaccine (CoronaVac) from China and first batch of vaccines arrived on 30 December 2020.

We collected Turkish contents related to vaccines published publicly on Twitter, between December 9, 2020 and January 8, 2021. Data collected via software developed by a researcher in Python programming language using open-source libraries and Twitter Application Programming Interface (API) (12). The software sends a request to Twitter using a search query and receives data of the content matching the query. The query used in this study is composed of the keyword “vaccine” (“aşı” in Turkish) and its derivatives. (Whole search query can be found in S1 Appendix). Due to restrictions of standard Twitter API, data collected on a weekly basis. In total 551,245 tweets were collected in the study period. To be able to grasp potential daily differences among contents, a sample was selected by stratified random sampling proportioned to the daily number of tweets. A sample size of 1000 tweets considered to be enough to reach data saturation. Eventually, 1041 tweets were included in study sample.

Data for following variables are collected for each tweet and their publishers: text, publication time, presence of a media, presence of an URL, presence of a hashtag; publisher’s duration of twitter use, number of followers, number of tweets, and account verification status.

Four researchers analyzed and coded 260 tweets independently. Researchers analyzed content in the following means: relevancy with the vaccine, type of the user (organizational or personal), how the vaccine is named (if available), the anti-vaccine argument in the content (if available). After completion of the analysis, data regarding to anti-vaccine argument codes were gathered, data saturation was affirmed, and emerging themes that are agreed on were identified by the researchers.

Emerging themes are reported with example quotations (original Turkish texts of quotations can be found in S2 Appendix). Additionally, descriptive statistics for independent variables are summarized in number and percentage for categorical variables; median and interquartile range for continuous variables that are not distributed normally.

## Results

### Descriptive statistics

Descriptive characteristics of the user accounts and the tweets are presented in Table 1 and Table 2 respectively. All tweets were published from 1000 unique accounts of which 2.7% were verified and 11.3% organizational users. Median duration of twitter use was 4.0 years, the median number of followers was 276.5 and median number of tweets was 3163.5 (Table 1).

**Table 1:** Characteristics of the user accounts.

|  | n | % |
| --- | --- | --- |
| **Verification status** |  |  |
| Not verified | 973 | 97.3 |
| Verified | 27 | 2.7 |
| **User type** |  |  |
| Personal or others | 887 | 88.7 |
| Organizational | 113 | 11.3 |
| **TOTAL** | **1000** | **100.0** |
| **Median (IQR)** |  |  |
| **Duration of Twitter use (year)** | 4.0 (1.0-8.0) |  |
| **Number of followers** | 276.5 (55.0-603.8) |  |
| **Number of tweets published** | 3163.5 (1561.5-13,951.0) |  |

**Table 2:**
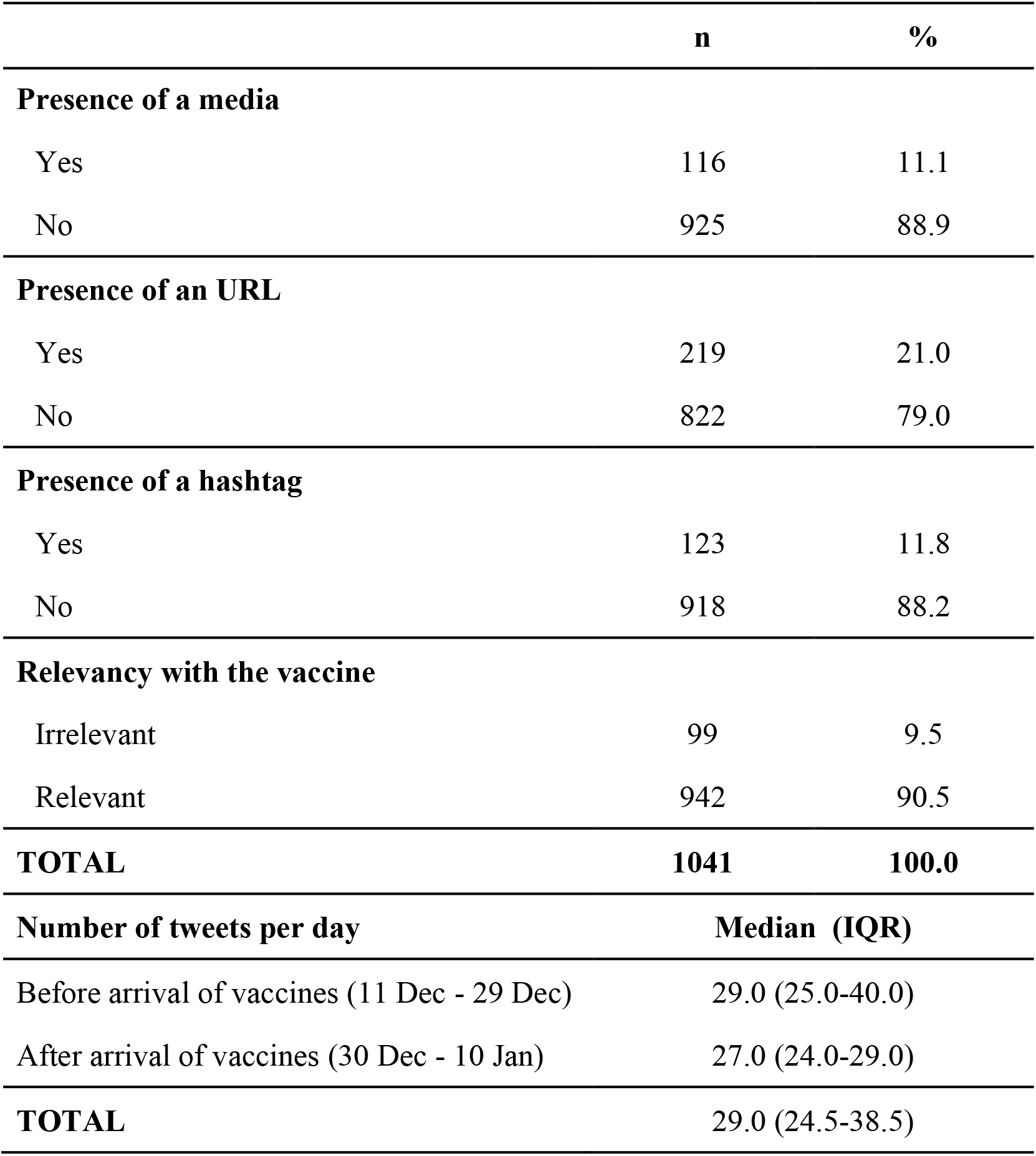
Characteristics of the tweets.

|  | **n** | **%** |
| --- | --- | --- |
| **Presence of a media** |  |  |
| Yes | 116 | 11.1 |
| No | 925 | 88.9 |
| **Presence of an URL** |  |  |
| Yes | 219 | 21.0 |
| No | 822 | 79.0 |
| **Presence of a hashtag** |  |  |
| Yes | 123 | 11.8 |
| No | 918 | 88.2 |
| **Relevancy with the vaccine** |  |  |
| Irrelevant | 99 | 9.5 |
| Relevant | 942 | 90.5 |
| **TOTAL** | **1041** | **100.0** |
| **Number of tweets per day** |  | **Median (IQR)** |
| Before arrival of vaccines (11 Dec - 29 Dec) |  | 29.0 (25.0-40.0) |
| After arrival of vaccines (30 Dec - 10 Jan) |  | 27.0 (24.0-29.0) |
| **TOTAL** |  | **29.0 (24.5-38.5)** |

Among 1041 tweets included in the study 90.5% were about vaccines, 11.1% included at least a media and 21.0% included URL. Median number of tweets per day was 29.0 during 11-29 Dec and 27.0 during 30 Dec-10 Jan (Table 2).

Daily numbers of tweets are shown in Fig 1. The median number of included tweets per day was 29.0 during 11-29 December which corresponds to the time before arrival of the first group of vaccines to Turkey.

**Fig 1.**
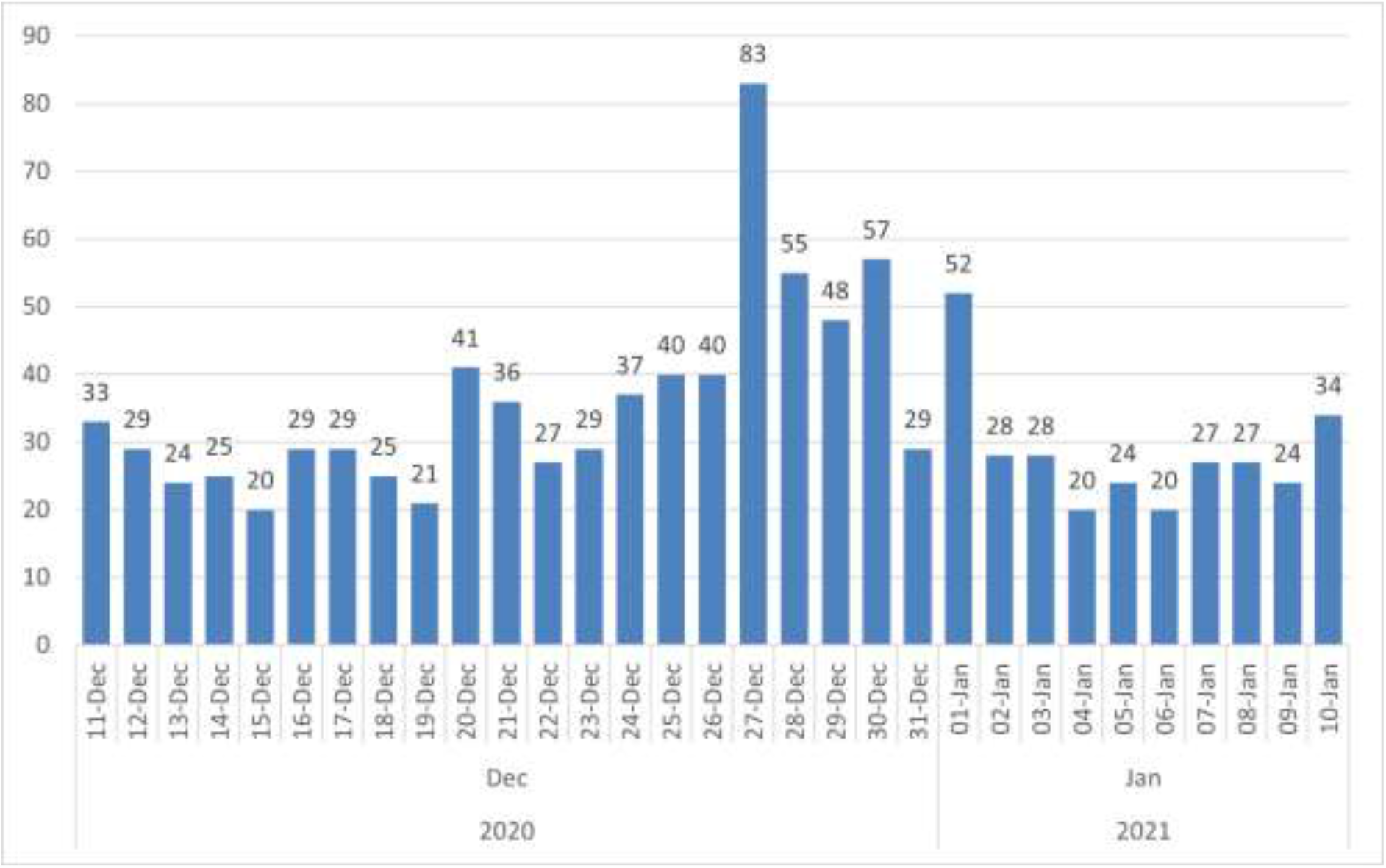
Daily numbers of tweets.

Content characteristics of the tweets are summarized in Table 3. As it is seen in Table 3, 22.6% (n=213) of the tweets included at least one name of a COVID-19 vaccine, 2.7% (n=25) other vaccine names and 74.8% (n=705) did not include any vaccine name. 22.0% (n=207) of the tweets included at least one anti-vaccination theme.

**Table 3:** Tweet contents including vaccine names and anti-vaccination themes.

| Contents | n | % |
| --- | --- | --- |
| **Vaccine names** |  |  |
| Name of a COVID-19 vaccine | 213 | 22.6 |
| Name of other vaccines* | 25 | 2.7 |
| No vaccine name | 705 | 74.8 |
| **TOTAL** | **942** | **100.0** |
| **Anti-vaccination themes** |  |  |
| Including tweets | 207 | 22.0 |
| Not including tweets | 735 | 78.0 |
| **TOTAL** | **942** | **100.0** |

\* Influenza, pneumonia, rabies, multiple sclerosis, tetanus, smallpox, polio, cholera, human papillomavirus. One tweet mentioned both COVID-19 and multiple sclerosis vaccines.

Frequency distribution of COVID-19 vaccine names and anti-vaccination themes are presented in Tables 4 and 5 respectively.

**Table 4:** Distribution of the vaccine names in tweets that mentioned a COVID-19 vaccine.

| **Names of the Vaccines** | **n\*** | **%** |
| --- | --- | --- |
| **CoronaVac mentions** | **109** | **51.2** |
| Chinese vaccine | 90 | 42.3 |
| Sinovac | 19 | 8.9 |
| CoronaVac | 4 | 1.9 |
| **Comirnaty mentions** | **57** | **26.7** |
| Pfizer-Biontech | 54 | 25.4 |
| German vaccine | 5 | 2.3 |
| Comirnaty | 1 | 0.5 |
| **Moderna mentions** | **14** | **6.6** |
| Moderna | 12 | 5.6 |
| American vaccine | 4 | 1.9 |
| **National (Turkish) vaccine mentions** | **12** | **5.6** |
| **mRNA Vaccine mentions** | **8** | **3.8** |
| **AstraZeneca mentions** | **7** | **3.3** |
| Oxford | 4 | 1.9 |
| AstraZeneca | 3 | 1.4 |
| **Sputnik V mentions** | **5** | **2.3** |
| Russian vaccine | 4 | 1.9 |
| Sputnik V | 1 | 0.5 |
| **Other COVID-19 Vaccine mentions** | **14** | **6.6** |
| **TOTAL** | **213** | **100.0** |

\* Sum of the numbers is not equal to the total and subtotals because some tweets included more than one vaccine name.

**Table 5:**
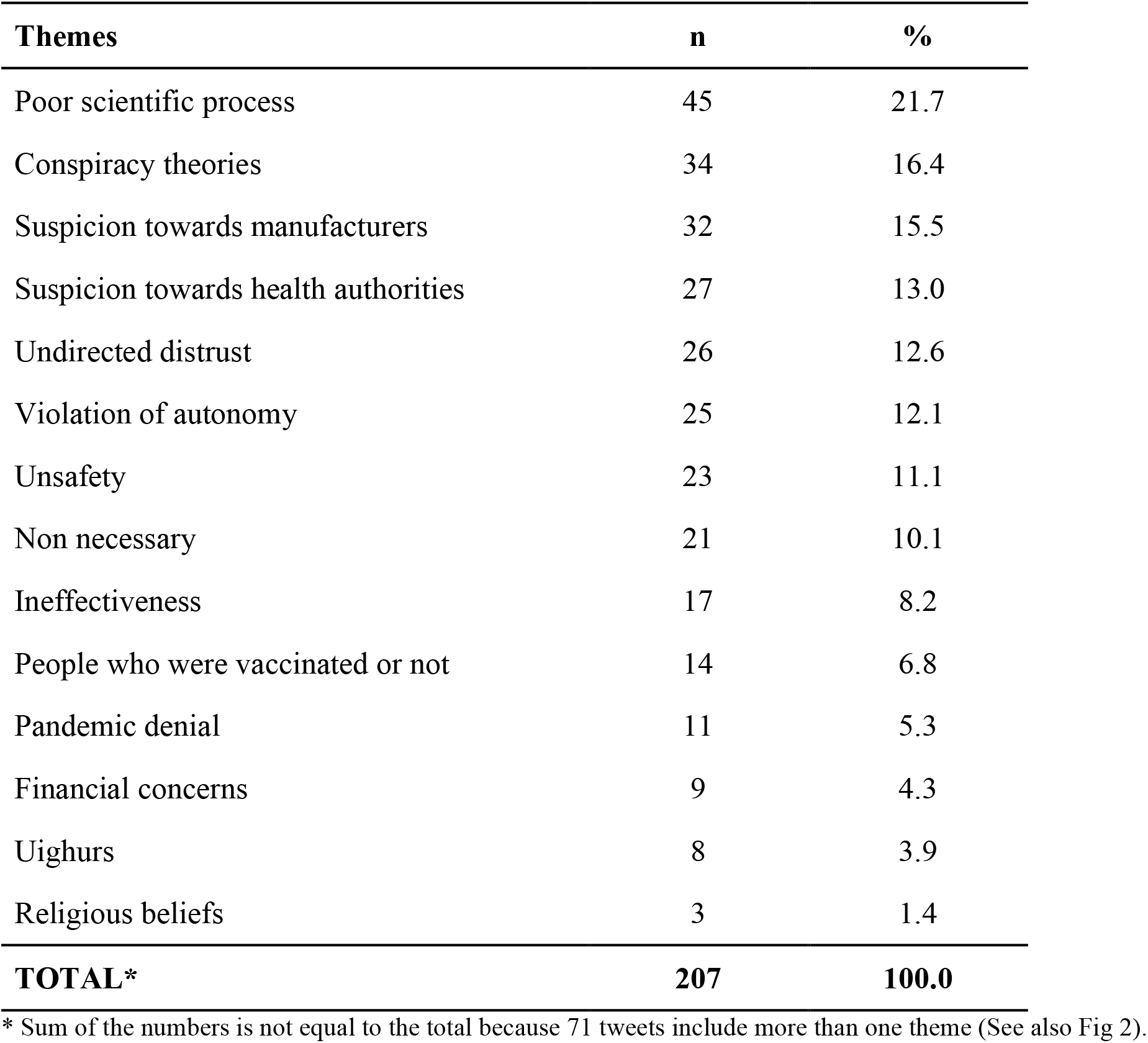
Distribution of anti-vaccination themes.

| Themes | n | % |
| --- | --- | --- |
| Poor scientific process | 45 | 21.7 |
| Conspiracy theories | 34 | 16.4 |
| Suspicion towards manufacturers | 32 | 15.5 |
| Suspicion towards health authorities | 27 | 13.0 |
| Undirected distrust | 26 | 12.6 |
| Violation of autonomy | 25 | 12.1 |
| Unsafety | 23 | 11.1 |
| Non necessary | 21 | 10.1 |
| Ineffectiveness | 17 | 8.2 |
| People who were vaccinated or not | 14 | 6.8 |
| Pandemic denial | 11 | 5.3 |
| Financial concerns | 9 | 4.3 |
| Uighurs | 8 | 3.9 |
| Religious beliefs | 3 | 1.4 |
| **TOTAL\*** | **207** | **100.0** |

\* Sum of the numbers is not equal to the total because 71 tweets include more than one theme (See also Fig 2).

As it is seen from Table 4, 213 tweets included a total of 235 COVID-19 vaccine names. Most frequently mentioned COVID-19 vaccine was CorronaVac (51.2%). Yet, it was mostly expressed as “Chinese vaccine” (42.3%).

### Emerging themes

Emerging anti-vaccination themes in the contents of the tweets are as listed below:

1. Poor scientific process
2. Conspiracy theories
3. Suspicion towards manufacturers
4. Suspicion towards health authorities
5. Undirected distrust
6. Violation of autonomy
7. Unsafety
8. Non necessary
9. Ineffectiveness
10. People who were vaccinated or not
11. Pandemic denial
12. Financial concerns
13. Uighurs
14. Religious beliefs

Among all tweets 22.0% (n=207) had an anti-vaccination theme. In total 207 tweets included 295 themes (56 tweets include two, 13 tweets include three and 2 tweets include four themes.). Frequency distribution of the themes are presented in Table 5

Among tweets that included an anti-vaccination theme; poor scientific processes (21.7%), conspiracy theories (16.4%), and suspicions towards manufacturers (15.5%) were the most frequently mentioned themes.

Seventy-one tweets (34,2%) were coded for more than one theme. Co-occurence of themes among those tweets is visualized in Fig 2. At most, “poor scientific process” theme come along with “suspicion towards manufacturers” (n=9) and “suspicion towards health authorities” (n=5).

**Fig 2.**
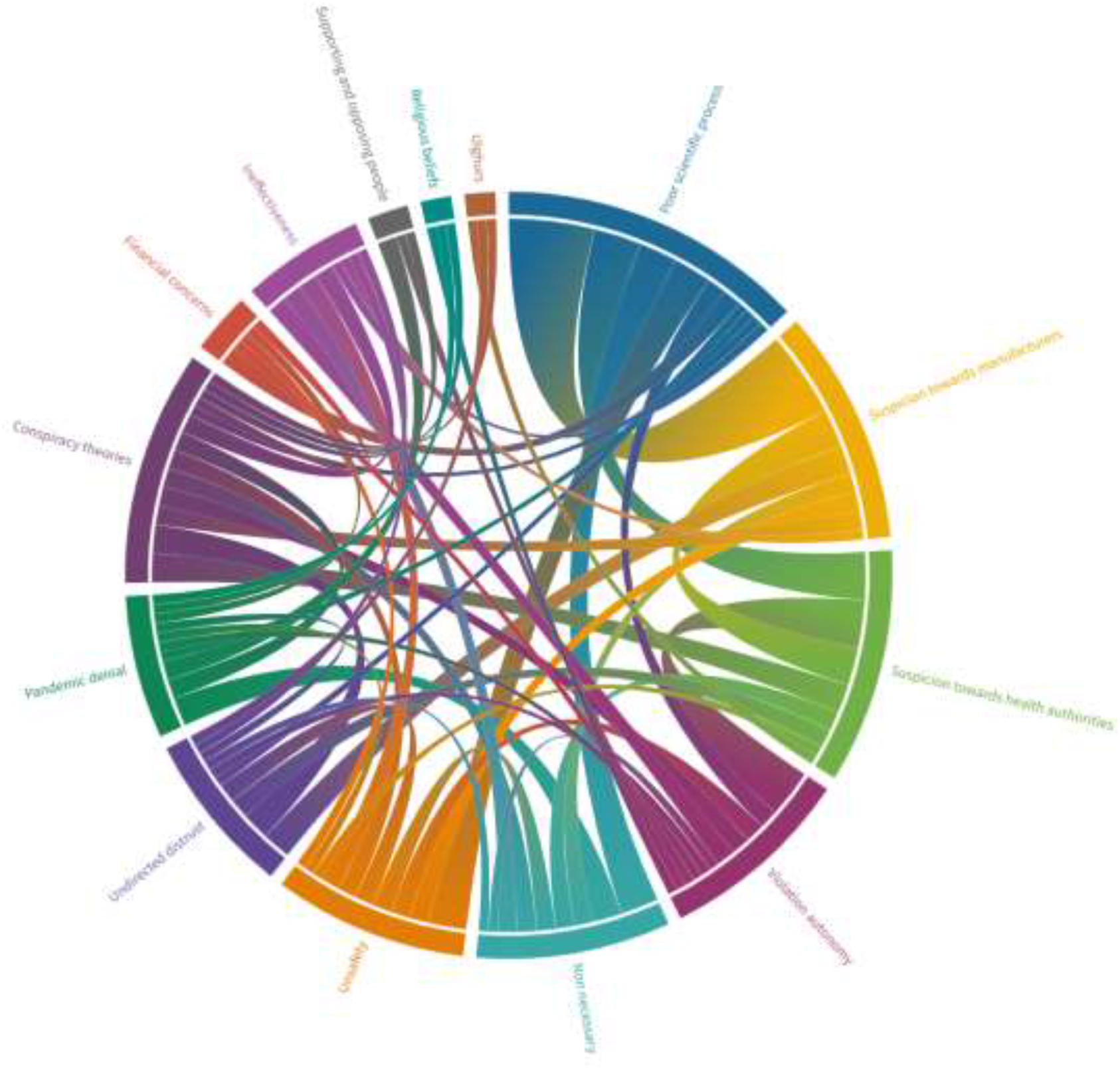
Co-occurrence of themes among tweets that contain more than one theme.

#### Poor scientific process

Suspicions about the scientific research processes in which vaccines are produced were the most frequent theme. Claims such as lack of sufficient scientific data on vaccines, the quick development procedure of vaccines, and humans are going to be experimental subjects for vaccines were the main arguments in these themes.

> *Example 1: Why do we get the vaccine that even [nation name] government does not use it for its own people? We are not experimental subjects*.
>
> *Example 2: The only way to end the pandemic is to get vaccinated. However, no scientific evidence has been presented regarding the efficacy and the safety of the current vaccines. Without safety, no vaccine should be given to large populations*.

#### Conspiracy theories

There were claims stating that pandemic is a set-up of some mysterious international powers, and that the vaccine is going to be used as a biological weapon to change the demographic structure.

> *Example 1: Do you really believe all of these? They all tell you what globalists order them. First, it was supposed to end in 2021 summer, but now we are in 2021 winter, and they already extend it for the next ten years: :D :D I think it will end in 2071! My final call. What about those saying that vaccine is the light at the end of the tunnel? Vaccines are useless apparently :D*
>
> *Example 2: This is only the beginning. If we ain’t gonna stop all these today, their next steps might be even more dangerous. Don’t you get that the real matter is not COVID, vaccine, 5G, nor mask!*

#### Suspicion towards manufacturers

This theme refers to the expressions of suspicions towards the vaccine producer companies or countries.

> *Example 1: While [nation name] gets the vaccines from [another nation name] for its citizens, they sell us their illegal vaccine that does not even complete phase 3*.
>
> *Example 2: Today, I’ve heard rumors about, “[company name] and [nation name] vaccines are produced with active and passive viruses, respectively. And those who’ve gotten the [company name] are miserable right now*.*” I don’t know about their accuracy and I could not see any news about them. But, as I said, it’s all getting chaotic because they do not have any kind of safety mechanism*.

#### Suspicion towards health authorities

Some tweets expressed suspicions against the credibility of health authorities. Dissatisfaction about countries’ methods of combating the pandemic and claims of secret relationships in the health sector are included in such tweets.

> *Example 1: [scientist name] [company name] [vaccine brand] etc. You can show the dirty past of all vaccine producers and medical companies. You can countin all the doctors and their institutions as well. And all the governments that took side… Media, don’t be afraid. #GLOBALDECEPTION*
>
> *Example 2: Dear minister, we don’t believe in you anymore. You’ve ruined the lives of all people and artisans. They are all #GLOBALDECEPTION We all are sick of your lies and do not trust in your vaccines. And, we are not gonna get any of them. [URL]*

#### Undirected distrust

There have been some statements that there are many uncertainties about vaccines, and that they cannot be trusted yet. There were claims stating that the vaccine is a lie, and actually it does not exist.

> *Example 1: Lies about pandemic and institutions that fuel the fear among society… Covid might be a game or more of a trailer for a bigger game. Even the vaccination is a huge mystery*.
>
> *Example 2: It should be all followed to see where this mutation stuff will be related to. I feel like it’s gonna be a reason for the mandatory vaccination instead of the second wave. Almost all of the vaccination companies have already stated that the vaccines are effective even when the virus has mutated*.

#### Violation of autonomy

Objections to the mandatory vaccination in terms of the privacy of human body were included in this theme. Among the arguments presented were the forced vaccination and the imposition of this decision by the national public authorities on behalf of the citizen.

> *Example 1: It is not obligated yet we are not happy with this vaccination imposition. #CitizenisAreTheState*
>
> *Example 2: So they all are gonna make debates through my body about the mandatory vaccination, but I am gonna shut up, not look it up and come into the line like a sheep. Is that so?*

#### Unsafety

Tweets claiming that the vaccine has various harms on the body are included in this theme. There are arguments that the vaccine has fatal effects, its side effects are severe, and it may even cause cancer.

> *Example 1: Nobody knows the side effects of that vaccine on me. I prefer not having a vaccine that may harm or kill me, just for the sake of protecting others. And we all have a right to do this, do you get that? The vaccine or the experiment that’s gonna happen to me, I’ll make the call for that. What do you expect?*
>
> *Example 2: If the vaccine will result in malformed births for the next generations*.. *I am pretty sure it will be*.. *(There are tons of examples for the malformed births in Africa and India because of the mRNA vaccines). In order to avoid its damages, the unvaccinated generation should not marry with the vaccinated generation. You can’t see the extent of its damages*..

#### Non necessary

The claims that vaccines are not necessary in combating pandemics were the contents of this theme. It was among the statements that the disease is mild, the mortality rate is low, and that strong natural immunity is sufficient.

> *Example 1: Did you even end the flu with vaccine? You cannot vaccinate a virus that’s mutated. All we need is to have a strong immune system. That’s it*.
>
> *Example 2: We do not wanna get vaccination or something like that. Does anybody hear our voice??? I’ve survived from corona at home without any medicine. My relative that had a hard time during their recoveries also survived from it at thome. The half of my country people have survived it. I don’t wanna get a vaccination for something that I’ve already gotten the immunity for*.

#### Ineffectiveness

The claims that vaccines are ineffective against the virus, and therefore against the pandemic has been frequently expressed. Among the statements were the possibility of getting sick despite the vaccine, that it did not work against mutant viruses, and did not prevent transmission and death.

> *Example 1: I think the vaccination is not the way to end the disease. Whats’ important is that we need to develop medicines that can lead people to survive from this disease easily. The flu vaccinations are not %100 protective and they’re never gonna be. Everybody gets caught a flu and gets well with a medicine. That’s the only solution that we should focus*.
>
> *Example 2: Does the COVID-19 vaccine not working well? A doctor who got the vaccine 6 days ago just caught the corona virus again*.

#### Supporting and opposing people

Statements were made on opinion leaders or celebrities who got vaccinated or declared that they would not. There were expressions such as the low number of people who stated that they would be vaccinated, the majority of those who stated that they would not, and the distrust of someone who endorse the vaccine.

> *Example 1: [Controversial celebrity name] praised the [nation name] vaccine and said that those who discredit it create negative perception. I hope you understand why we should question this vaccine*.
>
> *Example 2: Don’t fool yourself, none of them have gotten the vaccine because there is no such a thing as covid. You don’t wanna get this. Do you really think that they all get the vaccine?*

#### Pandemic denial

This theme included another frequent claim regarding the denial of the existence of the pandemic. There were claims that the disease or virus did not actually exist, it was a lie, and fake.

> *Example 1: Although it might be a regular vaccine, there is no need for it because there is no such a disease. But, how are you really gonna believe whether those people in the media saying that they got vaccinated? Also, its effects should last 4-5 years at least*.
>
> *Example 2: Look at these photos that were just taken*.. *Also in Wuhan… Look, how China is messing with you al*.. *Even there is a virus for those tribe countries that have almost no people living in them, China with 2 billion people is joking with the world. No vaccine, no treatment, yet we are done with virus, they says. WAKE UP PEOPLE THERE IS NO VIRUS*

#### Financial concerns

Under this theme, there were some contents that the vaccines emerged completely because of financial concerns. There were reasons such as the vaccine being commercial, expensive and paid, and the goal was to earn money.

> *Example 1: Those people used to say that vaccines will be free but now trying to make profit out of them. They’ll even get taxes*.
>
> *Example 2: They all are really trying to make profit out of it. At the beginning, they all said that vaccines will be free, and now there are rumors saying that one dose will be 10 dollars*.

#### Uighurs

A few tweets were about the relationship between China’s East Turkestan policies and the vaccination process. There were claims that vaccines were provided to Turkey by a country that also persecuted the Uighurs and a commercial relationship with this country would mean betrayal to them.

> *Example 1: Things have done to our Uighur Turkish brothers are never ending. Also, we are contributing to Chinese economy by taking the vaccination from them although the Chinese government is the responsible for all pressures and tortures. These vaccinations are betrayal to our Uighurian brothers #UighursCannotBeRepatriated*
>
> *Example 2: Let’s stop all the fuss and I am not gonna get a vaccine or not even let them into my apartment that gathers Uighurians into more than 500 different camps under the so-called a training program*

#### Religious beliefs

There were also those who objected to vaccinations on religious beliefs. These people claimed that the vaccines were not halal because of their ingredients.

> *Example 1: We are not against the vaccine. We are against the vaccines that contain haram stuff. We wanna get halal drug and halal vaccine. #Vaccine #Drug #Halal #Local #National*
>
> *Example 2: How’s it gonna be caiz both for them and for muslims? Whats the difference if we all gonna get that piggy foetus mRNA vaccine that gonna change our genetic codes? But, see the Pope does not even wear a mask. But of course it should be an exception because mask is a symbol of slavery and not gonna work out for them*.

## Discussion

Our study is the first twitter-based qualitative content analysis in Turkey on COVID-19 anti-vaccine and hesitancy. Another key point of the study is that it includes tweets on Twitter during the delivery of the first batch of vaccines to Turkey.

Among all vaccine relevant tweets, 22.6% of them mentioned a name of COVID-19 vaccine and 22.0% included at least one anti-vaccination theme.

CoronaVac was the most frequently mentioned vaccine mostly with the expression “Chinese vaccine”. Interestingly, people described this vaccine mostly by its country of origin. Comirnaty was the second one and mostly expressed as Pfizer-Biontech vaccine which is the company of origin. Other vaccines were mostly expressed by their company of origin, as well.

During the analysis of the anti-vaccination contents of the tweets, we identified fourteen major themes. “Poor scientific process” was the predominant theme and it was followed by “Conspiracy theories”, and “Suspicion towards manufacturers” themes.

“Poor scientific process” theme included tweets mainly focused on concerns such as the lack of sufficient scientific data, the rapid development of vaccines, and vaccines that would be tested on the community by seeing them as an experimental subject. We of the opinion that this theme frequency was higher than others because of the fast-track vaccine development process due to the emergency need of the vaccine to reduce the effects of the pandemic. Berry et al.’s study showed similar concern as “The vaccine was developed too quickly” (13). Also, lack of information about the results of phase 3 trials of Coronavac, which is the only COVID-19 vaccine that was delivered to Turkey during our research, may increase the arguments in this theme.

The second most frequent theme was “Conspiracy theories”. It included conspiracy theories such as the vaccine is developed to use as a biological weapon to change the global demographic structure. The arguments are similar to Sallam et. al’s online-based questionnaire conducted in Arab Countries (14) and Ortiz-Sánchez et al’s systematic review that was performed on certain databases to analyze networks’ information about the anti-vaccine movement (9). Sallam et al’s questionnaire results showed that 59.5% of the respondents believed that COVID-19 is a man-made virus and 40% of them thought that it made to force everyone to get the vaccine (14). According to Salali and Uysal’s research that was conducted in the United Kingdom (UK) and Turkey by an online survey, 18% in Turkey and 12% in the UK thought the origin of the virus was artificial (15). In our study, we found that 3.6% of the tweets had that kind of claim. Moreover, Nuzhath et al.’s content analysis that was conducted on Twitter showed similarities with our findings. That research also has a theme as “conspiracy theories” and it is the second most frequent theme like ours. According to their findings; this theme involves some theories such as “Vaccine is being developed to limit or control population size”, “Vaccine will contain Microchip or tracking device”, “5G/3G technology related to COVID-19 infection and vaccine”, and “Vaccine makers created COVID-19” that we also met under this theme in our study (16). A similar concern was also shown in Berry et al.’s study as “Microchip” (13).

The following themes’ main focus were “suspicion towards manufacturers” and “suspicion towards health authorities”. “Suspicion towards manufacturers” theme included the expressions of suspicions towards the companies or countries that produce the vaccine. “Suspicion towards health authorities” theme included dissatisfaction about countries’ methods of combating the pandemic and claims of secret relationships in the health sector. A similar determinant as the name “mistrust in health institutions” was found in a rapid literature review that included the vaccine confidence, trust, and hesitancy articles published between 2004 and 2014 in Europe (17).

Another frequent theme was “Violation autonomy” which involves objections to the obligation of vaccination in terms of human bodies as property. When a population is at risk such as the risk of the populations during a pandemic, collective interests come prior than individual ones. Also, some implementations such as quarantine, isolation, and social distancing, limit the individual’s freedom and autonomy (18). Under this condition, It may be one of the reasons, which provokes people to share their concern about mandatory vaccination.

Moreover, there were the theme “non necessary” which consisted of the disease was mild, the mortality rate was low, and that strong natural immunity was sufficient, and the theme “ineffective” that included the statements which were the possibility of getting sick despite the vaccine, the ineffectiveness against mutant viruses and the lack of the prevention transmission and death. These themes showed similarity with the determinants “not required” and “vaccines not effective” of the rapid literature review that was conducted in Europe before the COVID-19 pandemic (17). The “non necessary” theme is also similar to the Twitter-based research of Nuzhath et. al’s finding that immunization against coronavirus is not necessary due to COVID-19 infections resulted in low death rates (16).

Another frequent theme was “pandemic denial” which may negatively affect not only the vaccination but also the compliance to precautions such as keeping social distance and wearing masks.

According to previous studies in vaccine hesitancy and anti-vaccination areas, it was not a common concern that the origin of the country of a vaccine before. We found in a few tweets the theme “Uighurs” was more specific than other studies carried out in this area. We conclude that this theme has emerged from the vaccine Coronavac, which was usually expressed as “Chinese vaccine”, as a nationalist concern against China’s East Turkestan policies where Uighur people lived.

A number of potential limitations need to be considered in our research. We actually conducted the content analysis in Turkish then translated to English. The meaning of the original tweets and the misspellings may be lost in translation. Also, this research included tweets in a specific time period as during the delivery of the first batch of vaccines to Turkey. On the other hand, it doesn’t include the time during vaccination. Our research is also limited by our keywords. Since we collected the tweets by our keywords, we might not catch complex versions of the vaccine’s name. Finally, our research was limited to Twitter.

In conclusion, it is well known that vaccine hesitancy and anti-vaccination attitudes may negatively affect the population’s health, especially during a pandemic, and contents of the social media is an important source of early information about such attitudes. Analysis of the social media message contents may be helpful for health managers to identify the major issues and also to organize the preventive measures.

## Data Availability

Data cannot be shared publicly because of Twitter's data policy. Unique identifiers of tweets and researchers coding are available on request from the corresponding author.

## Supporting information

S1 Appendix. The search query.

S2 Appendix. Original texts of quotations.

(Contact the corresponding author for access to the files.)

